# Optimized testing strategy for the diagnosis of GAA-*FGF14* ataxia

**DOI:** 10.1101/2023.02.02.23285206

**Authors:** Céline Bonnet, David Pellerin, Virginie Roth, Guillemette Clément, Marion Wandzel, Laëtitia Lambert, Solène Frismand, Marian Douarinou, Anais Grosset, Ines Bekkour, Frédéric Weber, Florent Girardier, Clément Robin, Stéphanie Cacciatore, Myriam Bronner, Carine Pourié, Natacha Dreumont, Salomé Puisieux, Marie-Josée Dicaire, François Evoy, Marie-France Rioux, Armand Hocquel, Roberta La Piana, Matthis Synofzik, Henry Houlden, Matt C. Danzi, Stephan Zuchner, Bernard Brais, Mathilde Renaud

**Affiliations:** Laboratoire de Génétique, CHRU de Nancy, France; INSERM-U1256 NGERE, Université de Lorraine, Nancy, France; Department of Neuromuscular Disease, UCL Queen Square Institute of Neurology and The National Hospital for Neurology and Neurosurgery, University College London, London, United Kingdom; Department of Neurology and Neurosurgery, Montreal Neurological Hospital and Institute, McGill University, Montreal, QC, Canada; Service de Neurologie, CHRU de Nancy, France; Service de Génétique Clinique, CHRU de Nancy, France; Faculty of Medicine and Health Sciences, Université de Sherbrooke, Sherbrooke, QC, Canada; Department of Diagnostic Radiology, McGill University, Montreal, QC, Canada; Department of Neurodegenerative Diseases, Hertie-Institute for Clinical Brain Research and Center of Neurology, University of Tübingen, Tübingen, Germany; German Center for Neurodegenerative Diseases (DZNE), Tübingen, Germany; Dr. John T. Macdonald Foundation Department of Human Genetics and John P. Hussman Institute for Human Genomics, University of Miami Miller School of Medicine, Miami, FL, USA; Department of Human Genetics, McGill University, Montreal, QC, Canada

**Keywords:** spinocerebellar ataxia, late-onset ataxia, trinucleotide repeat expansion disorders, *FGF14*, genetic diagnosis, diagnostic protocol, long-range PCR, GAA-*FGF14* ataxia

## Abstract

**Background:** Dominantly inherited GAA repeat expansions in *FGF14* are a common cause of spinocerebellar ataxia (GAA-*FGF14* ataxia; SCA27B, late-onset). Molecular confirmation of *FGF14* GAA repeat expansions has thus far mostly relied on long-read sequencing, a technology that is not yet widely available in clinical laboratories.

**Methods:** We developed and validated a strategy to detect *FGF14* GAA repeat expansions using long-range PCR, bidirectional repeat-primed PCRs, and Sanger sequencing. We compared this strategy to targeted nanopore sequencing in a cohort of 22 French Canadian patients and next validated it in a cohort of 53 French index patients with unsolved ataxia.

**Results:** Diagnosis was accurately confirmed for all 22 French Canadian patients using this strategy. Method comparison showed that capillary electrophoresis of long-range PCR products significantly underestimated expansion sizes compared to nanopore sequencing (slope, 0.87 [95% CI, 0.81 to 0.93]; intercept, 14.58 [95% CI, -2.48 to 31.12]) and gel electrophoresis (slope, 0.84 [95% CI, 0.78 to 0.97]; intercept, 21.34 [95% CI, -27.66 to 40.22]). The latter techniques yielded similar size estimates. Following calibration with internal controls, expansion size estimates were similar between capillary electrophoresis and nanopore sequencing (slope: 0.98 [95% CI, 0.92 to 1.04]; intercept: 10.62 [95% CI, -7.49 to 27.71]), and gel electrophoresis (slope: 0.94 [95% CI, 0.88 to 1.09]; intercept: 18.81 [95% CI, -41.93 to 39.15]). We identified 9 French patients (9/53; 17%) and 2 of their relatives who carried an *FGF14* (GAA)_≥250_ expansion.

**Conclusion:** This novel strategy reliably detected and sized *FGF14* GAA expansions. It compared favorably to long-read sequencing and can readily be implemented in clinical laboratories.

## Introduction

The late-onset cerebellar ataxias (LOCAs) are a group of neurodegenerative conditions that have until recently largely challenged molecular diagnosis^1,2^. Despite recent advances in our understanding of the genetic basis of these conditions, a genetic diagnosis is reached in less than 50% of patients with LOCA^3,4^. Dominantly inherited GAA repeat expansions in the first intron of the Fibroblast Growth Factor 14 gene (*FGF14*) have recently been reported as a common cause of LOCA (GAA-*FGF14* ataxia; SCA27B, late-onset [MIM: 620174]), accounting for 10 to 61% of unsolved cases in various cohorts^5,6^. Current data support a pathogenic threshold of (GAA)_≥250_ repeat units. Core clinical features of GAA-*FGF14* ataxia include slowly progressive cerebellar ataxia, early episodic symptoms, downbeat nystagmus, diplopia, and dizziness/vertigo^5^. Molecular confirmation of the *FGF14* GAA repeat expansion has thus far mostly relied on long-read sequencing, a technology that is not yet widely available in clinical diagnostic laboratories. Given the high reported frequency of GAA-*FGF14* ataxia, there is an immediate need to establish a standardized, accessible and validated molecular strategy for diagnosing this novel repeat expansion disorder in clinical diagnostic laboratories.

Herein, we propose a strategy that combines long-range polymerase chain reaction (PCR), bidirectional repeat-primed PCR (RP-PCR) and Sanger sequencing to detect and resolve *FGF14* GAA repeat expansions in clinical diagnostic settings. We further demonstrate the applicability of this diagnostic approach in a cohort of French patients with unsolved LOCA.

## Methods

### Patient enrollment

All participants provided written informed consent. This study was conducted in accordance with the Declaration of Helsinki.

#### French Canadian cohort

Patients were recruited at the Montreal Neurological Hospital of the McGill University Health Centre (Montreal, QC, Canada). The institutional review board of the McGill University Health Centre approved this study (MPE-CUSM-15-915). The French Canadian participants included 22 patients with GAA-*FGF14* ataxia and six controls that were reported previously^5^. All participants underwent genotyping of the *FGF14* repeat locus by agarose gel electrophoresis of long-range PCR (LR-PCR) amplification products and targeted long-read nanopore sequencing, as described previously^5^.

#### French cohort

Fifty-three index patients with LOCA and two affected relatives were recruited at the Centre Hospitalier Régional Universitaire de Nancy (France). The institutional review board of the Centre Hospitalier Régional Universitaire de Nancy approved this study (2020PI220). Patients were enrolled if meeting the following inclusion criteria: (a) progressive ataxia with onset at or after age 30; (b) no clinical features suggestive of multiple system atrophy (MSA); (c) exclusion of acquired causes; and (d) exclusion of known genetic causes by testing negative on an ataxia gene panel and for common coding and non-coding repeat expansion disorders. The Scale for the Assessment and Rating of Ataxia (SARA)^7^ was recorded when possible. Magnetic resonance imaging (MRI) of the brain was obtained for nine patients with GAA-*FGF14* ataxia.

### Molecular analysis (Figures 1 and S1)

#### First step: Fluorescent long-range PCR

The intronic *FGF14* GAA repeat locus was amplified by fluorescent long-range PCR (fLR-PCR). fLR-PCR products were analyzed on an ABI 3130*xl* DNA Analyzer (Applied Biosystems, Foster City, CA, USA) using the GeneScan 1200 Liz Dye Size Standard (catalog no. 4379950, Applied Biosystems). Results were analyzed using the GeneMapper software (version 6.0, Applied Biosystems). The PCR primers are predicted to amplify a 300 bp fragment based on the reference sequence, which includes 50 GAA triplets. The size of the expansion was calculated using the following formula: (size of PCR amplification product - 150)/3.

#### Second step: Bidirectional RP-PCRs

Two RP-PCRs targeting the 5’ end (5’ RP-PCR) and the 3’ end (3’ RP-PCR) of the locus were used in parallel to ascertain the presence of a GAA expansion at the repeat locus. RP-PCR products were analyzed on an ABI 3130*xl* DNA Analyzer using the GeneScan 1200 Liz Dye Size Standard. Results were analyzed using the GeneMapper software. The presence of characteristic saw-toothed products indicated the presence of a GAA repeat expansion at the *FGF14* repeat locus.

#### Third step: Gel electrophoresis of long-range PCR products and Sanger sequencing

##### 1) Samples with a single normal allele detected on fLR-PCR

Cases with a single normal allele detected by capillary electrophoresis of fLR-PCR products, regardless of their RP-PCRs profiles, next underwent gel electrophoresis of LR-PCR amplification products using a 1.5% agarose gel. This step allows for distinguishing cases homozygous for two normal alleles from cases heterozygous for one normal allele and one expanded allele that is too large (>400 repeat units, see Results) to be detected by capillary electrophoresis. The motif of any expansion identified at this stage is determined by the RP-PCR profiles generated during step 2, with sawtooth profiles indicating GAA expansions and flat profiles indicating non-GAA expansions. Amplification products were migrated for 3h30 at 150V and 300 mA.

##### 2) Samples with at least one allele of 250 or more repeat units detected on fLR-PCR and no sawtooth profile or interrupted profile on RP-PCRs

Cases with at least one allele of 250 or more repeat units and no sawtooth profile or an interrupted profile on RP-PCRs next underwent Sanger sequencing to determine the sequence and repeat motif of the *FGF14* locus. Sanger sequencing of LR-PCR amplification products was performed using the Applied Biosystems 3130*xl* DNA Analyzer. The resulting sequences were analyzed using Sequence Scanner version 1.0 software (Applied Biosystems).

The primer sequences and experimental conditions are provided in Supplementary Table S1.

### Statistical analysis

We compared methods against each other using the Passing-Bablok regression model (R package: mcr). Confidence interval of the slope that does not include 1 indicate statistically significant evidence of a proportional bias between two methods. Confidence interval of the intercept that does not include 0 indicate statistically significant evidence of a systematic bias between two methods. Linear relationship between two sets of measurements was tested by the CUSUM test of linearity. We used the Bland-Altman analysis to compare the measurements of the same variable by two methods. Correlations were calculated using the Pearson correlation coefficient. We analyzed the data in R (version 4.1) and GraphPad Prism 9. *P* value of <0.05 was considered significant. All analyses were two-tailed.

## Results

### *FGF14* repeat sizing by fLR-PCR

We first determined how repeat size estimates of larger alleles by fLR-PCR compare and correlate to (1) targeted long-read nanopore sequencing and (2) LR-PCR estimates by agarose gel electrophoresis estimates. We used 28 French Canadian participants for whom long-read sequencing data was available and measured repeat length by fLR-PCR. Nanopore sizing was performed on size-selected alleles of more than 80 repeat units to increase coverage depth, as described previously^5^.

We observed that expansions (GAA)_>400_ repeat units could not be accurately sized by fLR-PCR, as shown in Figure 2A, as they fall beyond the limit of detection of capillary electrophoresis. Therefore, such large expansions needed to be sized by agarose gel electrophoresis of LR-PCR amplification products.

**Figure 1:**
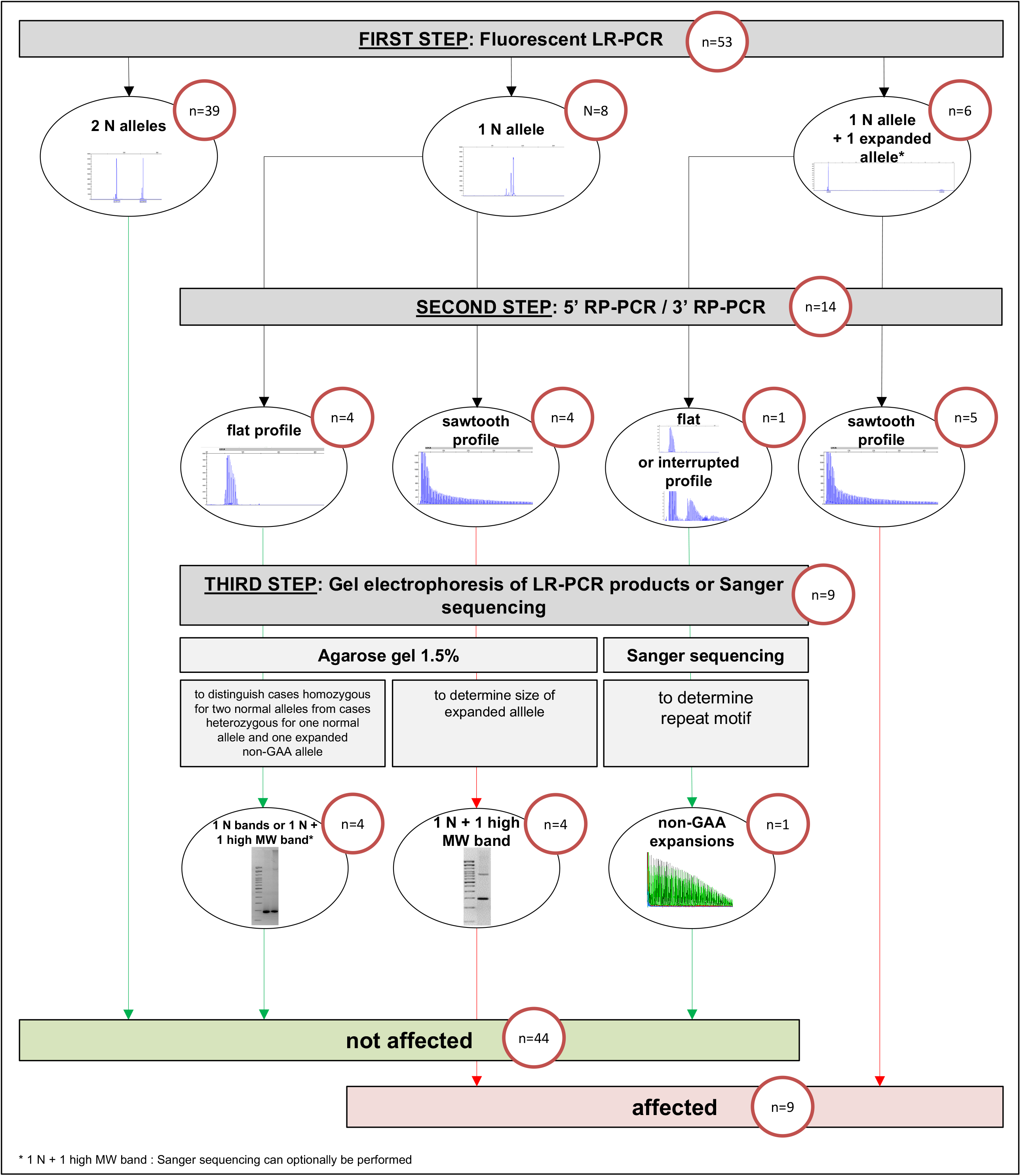
Testing strategy for the diagnosis of GAA-*FGF14* ataxia. The number of samples in the French Cohort of 53 index patients processed at each step is indicated in the red circles. Normal alleles have (GAA)_<250_ repeats and expanded alleles have (GAA)_≥250_ repeats. *Two expanded alleles are possible Legend: N allele, normal allele <250 repeat units; LR-PCR, long-range polymerase chain reaction; MW, molecular weight.

**Figure 2:**
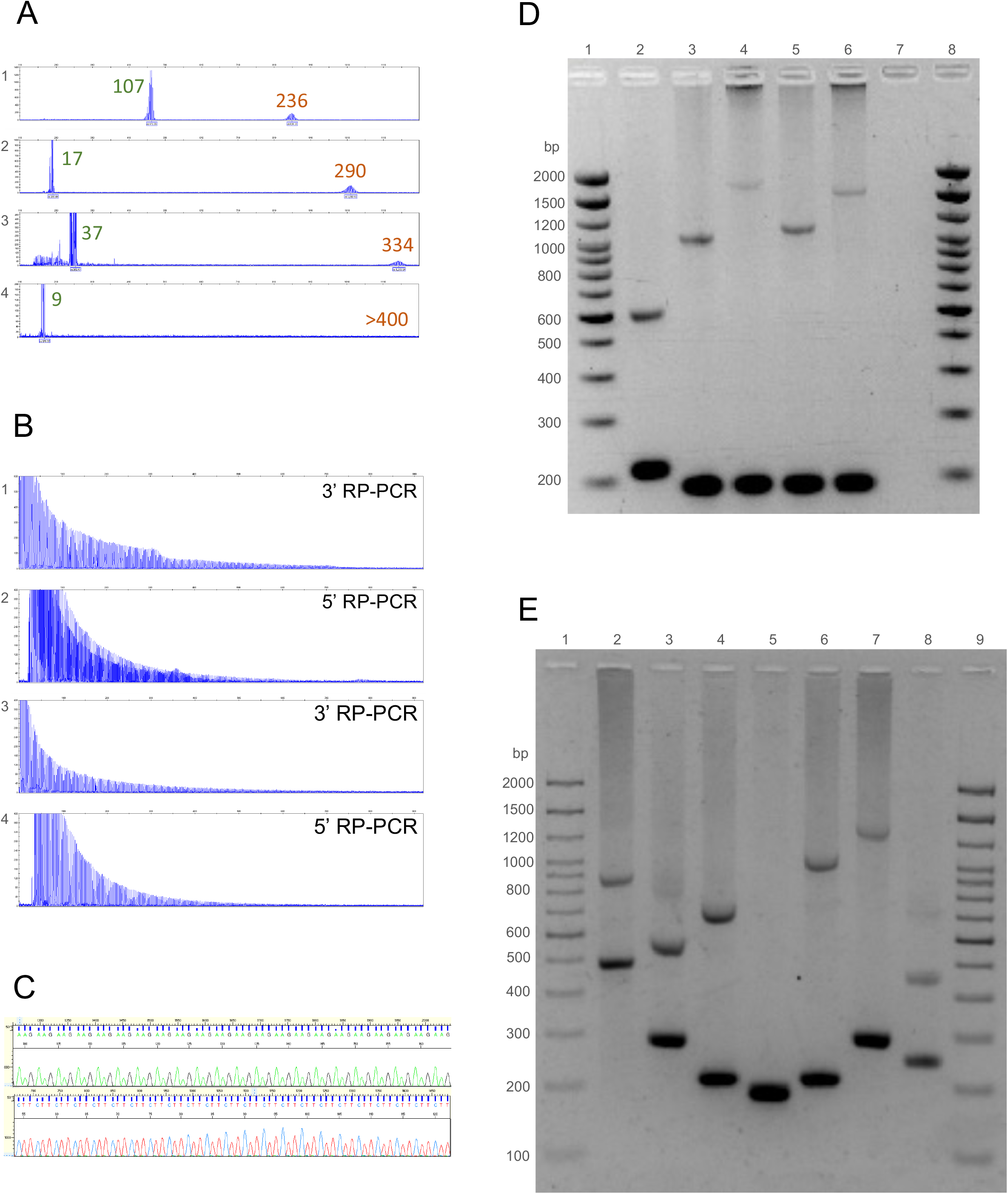
Molecular analysis of the *FGF14* repeat locus. (A) Fluorescent long-range PCR of the *FGF14* repeat locus of four patients with late-onset cerebellar ataxia; The calculated number of repeat units of each allele (before correction) is indicated for each of the four patients. (B) 5’ RP-PCR and 3’RP-PCR of the *FGF14* repeat locus of two patients carrying a (GAA)_≥250_ repeat expansion in *FGF14*. (C) Sanger sequencing of a patient carrying a (GAA)_≥250_ repeat expansion showing GAA repeats: (GAA)_n_ on forward strand and (TTC)_n_ on reverse strand. (D) Agarose gel electrophoresis (1.5%), lanes 1 and 8: 2,000 bp molecular weight marker, 2: 17/132 repeat units, 3: 9/280 repeat units, 4: 9/510 repeat units, 5: 9/313 repeat units, 6: 9/467 repeat units; 7: negative control. (E) Agarose gel electrophoresis (1.5%), lanes 1 and 9: 2,000 bp molecular weight marker, 2: 107/259 repeat units, 3: 40/132 repeat units, 4: 16/182 repeat units, 5: 8/9 repeat units, 6: 17/319 repeat units; 7: 43/370 repeat units, 8: 28/94 repeat units.

Consistent with what has previously been reported with other trinucleotide repeat expansions^8–12^, we observed a significant discrepancy between sizes of larger alleles measured by capillary electrophoresis and long-read sequencing or agarose gel electrophoresis. We compared methods using the Passing-Bablok regression model^13^, and found a statistically significant proportional bias between nanopore sequencing and fLR-PCR (slope, 0.87 [95% CI, 0.81 to 0.93] and intercept, 14.58 [95% CI, -2.48 to 31.12]) and gel electrophoresis and fLR-PCR (slope, 0.84 [95% CI, 0.78 to 0.97] and intercept, 21.34 [95% CI, -27.66 to 40.22]) (Figure 3A,B). Bland-Altman analysis confirmed a negative bias of -8.54% (95% limits of agreement, -13.34% to -3.75%) between fLR-PCR and nanopore sequencing, and of -10.91% (95% limits of agreement, -18.93 to -2.90%) between fLR-PCR and gel electrophoresis (Figure 3C,D). In comparison, the latter techniques yielded comparable size estimates (slope, 1.01 [95% CI, 0.90 to 1.11] and intercept, 5.35 [95% CI, -29.98 to 44.57]) (Figure S2A). Bland-Altman analysis of both techniques showed a positive bias of +2.55% (95% limits of agreement, -6.53% to 11.63%) (Figure S2B). These results confirm that capillary electrophoresis systematically underestimates size of expanded alleles compared to nanopore sequencing and gel electrophoresis, and suggest that GAA repeat-containing DNA fragments migrate faster than predicted. Faster migration of triplet repeat-containing fragments is thought to result from altered electrophoretic properties of repetition-rich DNA^11,12^. The systematic underestimation by capillary electrophoresis resulted in a false negative result in four of 28 participants (14%) of the French Canadian cohort carrying an *FGF14* expansion. To compensate for this underestimation, we used a set of four alleles whose sizes have been established by long-read sequencing and applied the least squares method to generate a straight line that fits our data and calculated its slope and y-intercept (Table S2). Application of this correction resulted in expansion size estimates being comparable between nanopore sequencing and fLR-PCR (slope, 0.98 [95% CI, 0.92 to 1.04] and intercept, 10.62 [95% CI, -7.49 to 27.71]) and gel electrophoresis and fLR-PCR (slope, 0.94 [95% CI, 0.88 to 1.09] and intercept, 18.81 [95% CI, -41.93 to 39.15]), as assessed by the Passing-Bablok regression model (Figure 3E,F). Bland-Altman analysis showed a positive bias of +1.23% (95% limits of agreement, -3.52% to +5.98%) between corrected fLR-PCR and nanopore sequencing, and a negative bias of -1.27% (95% limits of agreement, -9.16% to +6.61%) between corrected fLR-PCR and gel electrophoresis (Figure 3G,H). Allele sizing by fLR-PCR was highly reproducible, with an inter-assay variability of less than 0.3%.

**Figure 3:**
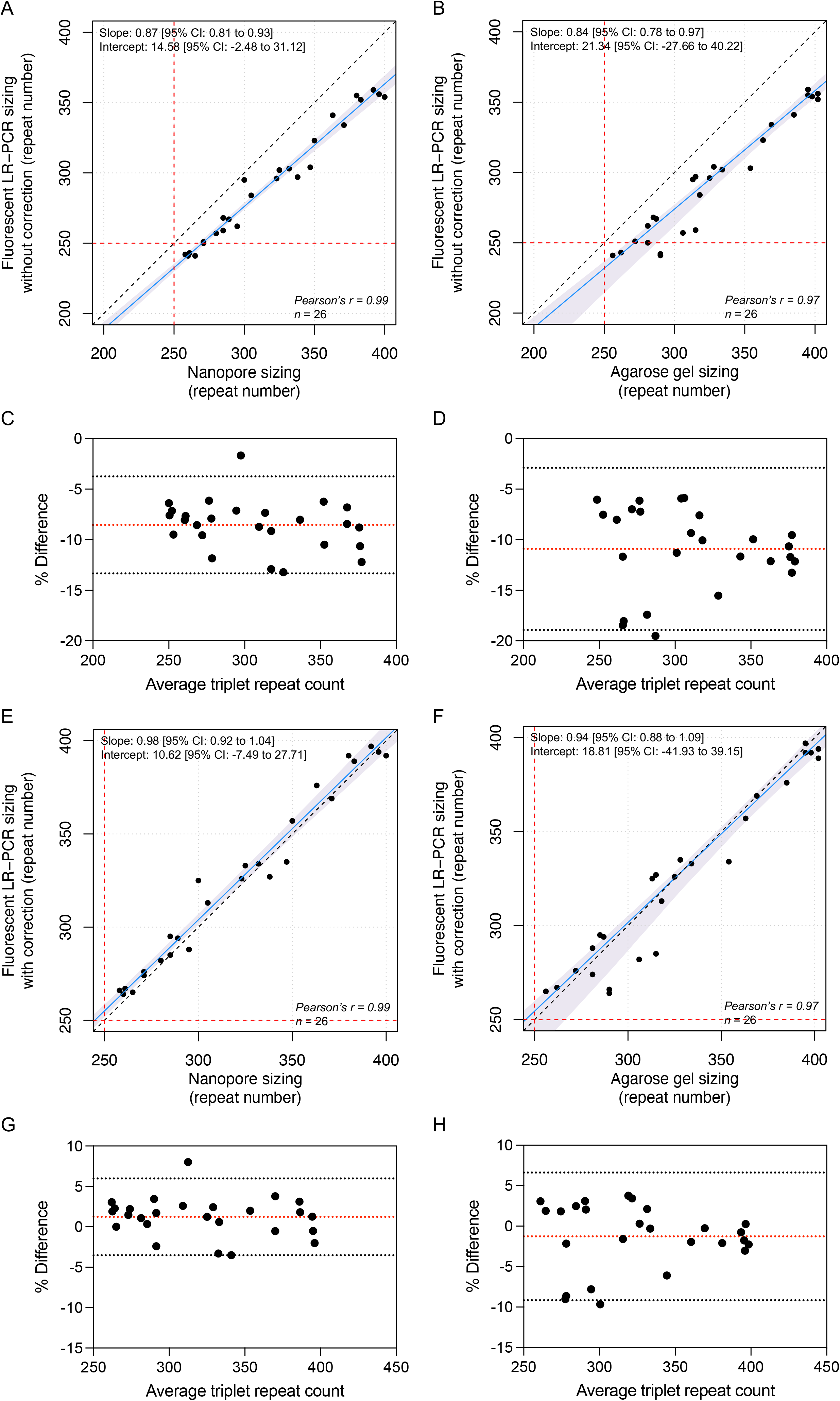
*FGF14* allele size estimates by fluorescent LR-PCR, long-read nanopore sequencing, and agarose gel electrophoresis. Passing-Bablok regression (blue lines) with 95% confidence interval (shaded blue areas) for allele size measured by (A) fLR-PCR and nanopore sequencing, (B) fLR-PCR and gel electrophoresis, (E) fLR-PCR (with correction) and nanopore sequencing, and (F) fLR-PCR (with correction) and gel electrophoresis. The dashed black lines show the identity line and the dashed red lines show the pathogenic threshold of (GAA)_≥250_ repeats. Bland-Altman plots show the percentage difference between size estimates measured by fLR-PCR and (C) targeted nanopore sequencing or (D) agarose gel electrophoresis as a function of the average of the two measurements for each sample. Plots show the percentage difference between size estimates measured by corrected fLR-PCR and (G) targeted nanopore sequencing or (H) agarose gel electrophoresis as a function of the average of the two measurements for each sample. The dashed red lines show the mean bias between two techniques and the dashed gray lines show the limits of agreement, defined as the mean percentage difference ± 1.96SD.

### Validation of the diagnostic approach for detection of *FGF14* GAA repeat expansions (Figures 1, 2, and S1)

The variability of the methods used thus far to resolve *FGF14* expansions^5,6^ and the high frequency of GAA-*FGF14* ataxia highlight the need to develop a standardized diagnostic approach that will be accessible and easy to implement in clinical diagnostic laboratories. Such approach must allow for accurate allele sizing, and assessment of repeat motif and sequence interruptions, if any. The former point is particularly important given that (GAA)_250-300_ alleles appear incompletely penetrant^5,6^. To address this need, we developed and validated a three-step strategy combining (1) fLR-PCR, (2) bidirectional RP-PCRs, and (3) agarose gel electrophoresis of LR-PCR amplification products and Sanger sequencing.

The first step of our proposed strategy involves fLR-PCR amplification of the *FGF14* repeat locus. The identification of two alleles of less than 250 repeat units ends the diagnostic process while any other pattern triggers additional testing. That includes samples with a single normal allele detected on fLR-PCR, since they may carry an expansion larger than 400 repeat units that falls beyond the limit of detection of capillary electrophoresis.

The second step involves bidirectional RP-PCRs targeting the GAA repeat unit at the 5’ end and the 3’ end of the locus. Bidirectional RP-PCRs allow for comprehensive assessment of the repeat motif over the entire length of even the larger expansions, and of sequence interruptions and polymorphisms at both ends of the locus. The identification of characteristic saw-toothed products, indicative of a GAA repeat expansion, in a sample found to have at least one allele of 250 or more repeat units on fLR-PCR confirms the diagnosis of GAA-*FGF14* ataxia and ends the diagnostic process. All other samples are moved to the third step.

As part of the third step, gel electrophoresis of LR-PCR products and Sanger sequencing are used to further resolve remaining samples. First, gel electrophoresis of LR-PCR products is performed on a 1.5% agarose gel for samples with a single allele detected by fLR-PCR, regardless of their RP-PCR profiles. This step allows for distinguishing cases homozygous for two normal alleles from cases heterozygous for one normal allele and one expanded allele that is otherwise too large to be detected by capillary electrophoresis. Gel electrophoresis additionally allows sizing of large expansions. The motif of any expansion is determined by the RP-PCR profiles generated during step 2, with sawtooth profiles indicating GAA expansions and flat profiles indicating non-GAA expansions. In case of non-GAA expansions, Sanger sequencing can optionally be performed to determine the repeat motif of the expansion. Second, Sanger sequencing is performed on samples with at least one allele of 250 or more repeat units measured by capillary electrophoresis and a flat or atypical profile on RP-PCRs – indicative of a non-GAA or interrupted expansion – to determine the repeat motif of the *FGF14* locus.

We validated this diagnostic strategy in a cohort of 53 index patients with unsolved LOCA that were recruited in Nancy, France. Using this strategy, we identified nine patients (17%) who carried (GAA)_≥250_ repeat expansions in *FGF14*. The expansion was also present in two affected relatives of one of the index patients. We also identified a patient compound heterozygous for expansions of 266 and 417 repeat units (Figure 4A: lane 3). RP-PCRs accurately determined the repeat motif (GAA and non-GAA expansions) of all expanded alleles in the cohort, as confirmed by forward and reverse Sanger sequencing. While Sanger sequencing of each individual strand fails to completely sequence expanded alleles, their combination allows for full sequence coverage of alleles of up to (GAA)_500_ repeat units.

**Figure 4:**
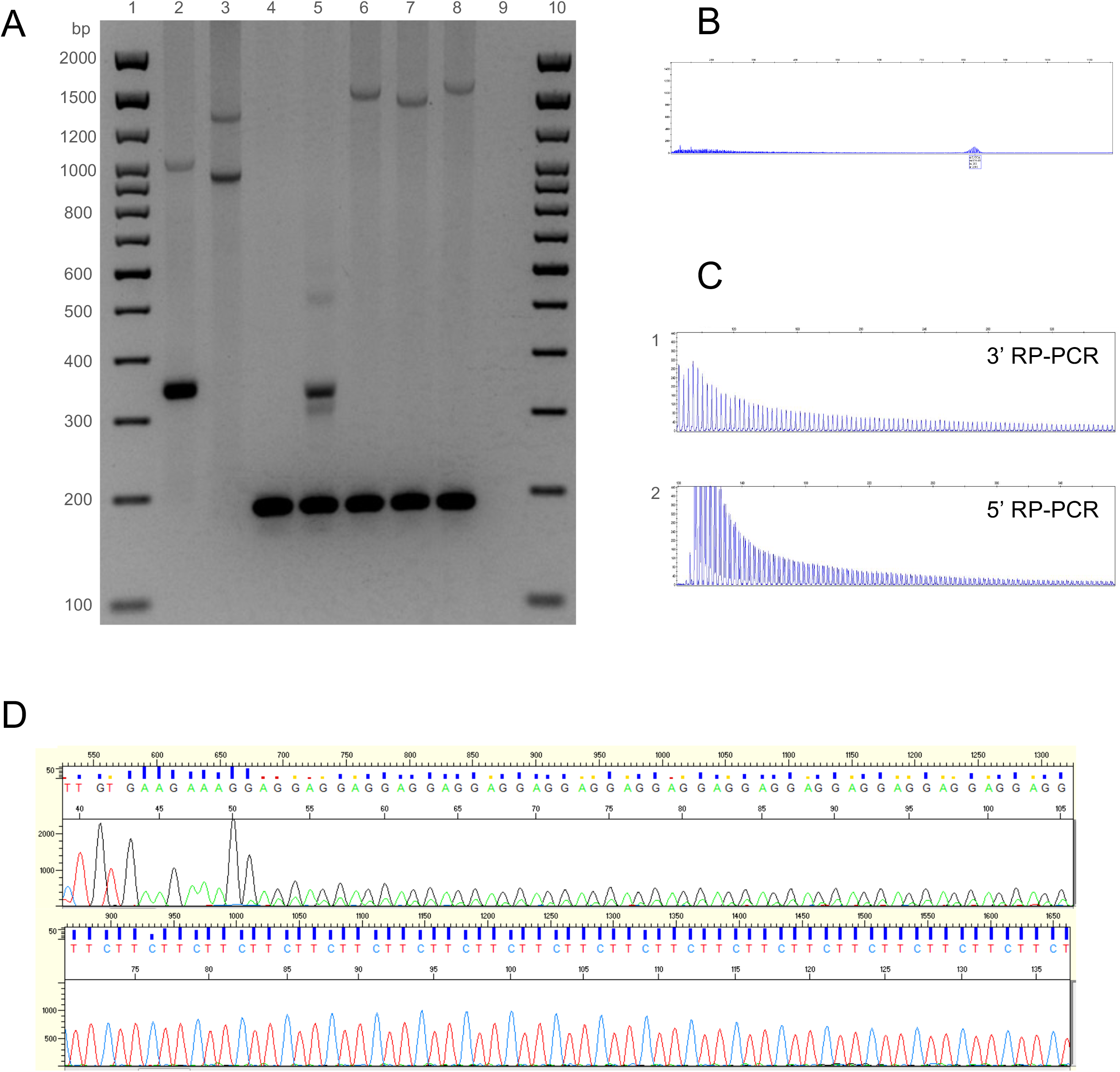
Molecular analysis of the *FGF14* repeat locus in a family with GAA-*FGF14* ataxia and a patient compound heterozygous for two expansions. (A) Agarose gel (1.5%), lanes 1 and 10: 2,000 bp molecular weight marker, 2: 58/300 repeat units, 3: 266/417 repeat units, 4: 9/9 repeat units, 5: 9/50 repeat units, 6: 9/492 repeat units, 7: 9/467 repeat units, 8: 9/510 repeat units, 9: negative control. The transmission of an expanded (GAA)_467_ allele resulted in expansion in the female germline in two meiotic events (expansion from 467 triplets to 492 and 510 triplets), as shown in lanes 7, 6 and 8, respectively. (B) Fluorescent long-range PCR, (C) 5’ RP-PCR and 3’RP-PCR, and (D) Sanger sequencing of a patient compound heterozygous for two expanded alleles (266/417 repeat units). (B) On fLR-PCR, the smallest expansion of 266 repeat units is detected whereas the larger expansion of 417 repeat units is not as it falls beyond the limit of detection of capillary electrophoresis. (D) Sanger sequencing shows polymorphism at the 5’ end of each expansion. Allele 1: (GAA)(GAAA)(GGA)(GAA)_n_ and allele 2: (GAA)(GAAA)(GAAA)(GAA)_n_.

Our diagnostic approach also allowed for the precise characterization of the sequence of each allele in the compound heterozygous patient (Figure 4A: lane 3, Figure 4B-D). While the 3’RP-PCR showed a typical sawtooth pattern, the 5’RP-PCR showed the superposition of two sawtooth profiles shifted by one base pair (Figure 4C). These profiles suggested that both GAA repeats were out of phase, as a result of the presence of an insertion or a deletion at the 5’ end of one allele. Sanger sequencing confirmed this hypothesis by showing the presence of a [(GAA)(GAAA)(GGA)] motif at the 5’ end of one allele and a [(GAA)(GAAA)(GAAA)] motif at the 5’ end of the second allele causing both alleles to be out of phase by one base pair.

### Clinical findings

The main clinical features of the French Canadian and French patients with GAA-*FGF14* ataxia are shown in Table 1. Early episodic features, nystagmus (gaze-evoked horizontal and downbeat) and alcohol intolerance were reported with similar frequency in patients from both cohorts. Episodic symptoms were reported in 73% of the French Canadian and French patients. The mean (±SD) age at the onset of episodic symptoms was 54±9 years and 64±9 years in the French Canadian and the French patients, respectively. The mean age at the onset of progressive ataxia was 59±10 and 66±10 years, respectively. Nystagmus was observed in 91% and 73% of patients, respectively. More than half of patients reported that alcohol intake triggered episodes of ataxia or markedly worsened baseline ataxia. At the chronic stage, the phenotype was characterized by a slowly progressive pan-cerebellar syndrome with predominant midline involvement. Gait ataxia was observed in 95% and 100% of French Canadian and French patients, respectively. Appendicular ataxia was observed in 90% of French Canadian and 91% of French patients. Pyramidal and extrapyramidal features were less frequently observed (each feature < 20% in the French Canadian cohort and French cohorts). Brain MRI showed mild to moderate cerebellar atrophy, with more severe involvement of the vermis, in 60% and 44% of patients, respectively. We observed an inverse correlation between the size of the repeat expansion and the age at disease onset (22 patients; Pearson correlation coefficient, -0.43; R^2^=0.18; *p*=0.048) or the age at onset of ataxia (22 patients, Pearson correlation coefficient, -0.55; R^2^=0.30; *p*=0.008) in the French Canadian cohort. In comparison, the size of the repeat expansion was not significantly associated with age at disease onset (11 patients; Pearson correlation coefficient, 0.07; *p*=0.85) or the age at onset of ataxia (8 patients, Pearson correlation coefficient, 0.16; *p*=0.71) in the French cohort.

**Table 1:** Characteristics of the French Canadian and French cohorts

|  | **French Canadian cohort**<br>**(n=22)** | **French cohort**<br>**(n=11)** |
| --- | --- | --- |
| *Male sex, no. (%)* | 13 (59%) | 4 (36%) |
| *Repeat count of GAA expansion, median (IQR)* | 333 (287-392) | 370 (313-492) |
| *Age at onset of episodic symptoms (mean ± SD)* | 54 ± 9 | 64 ± 9 |
| *Age at onset of permanent ataxia (mean ± SD)* | 59 ± 10 | 66 ± 10 |
| ***Inheritance*** |  |  |
| *Familial, no. (%)* | 17 (77%) | 7 (64%) |
| *Sporadic, no. (%)* | 5 (23%) | 3 (36%) |
| *Episodic onset, no. (%)* | 16 (73%) | 8 (73%) |
| *Alcohol intolerance, no. (%)* | 9/17 (53%) | 5/8 (63%) |
| *Nystagmus (gaze-evoked horizontal and downbeat), no. (%)* | 20 (91%) | 8 (73%) |
| *Episodic diplopia or visual blurring, no. (%)* | 15/21 (71%) | 5/9 (56%) |
| *Gait ataxia, no. (%)* | 21 (95%) | 10 (100%) |
| *Appendicular ataxia, no. (%)* | 19/21 (90%) | 9 (91%) |
| *Cerebellar dysarthria, no. (%)* | 8/21 (38%) | 7 (64%) |
| *Vertigo or dizziness, no. (%)* | 8/20 (40%) | 5/10 (50%) |
| *Cerebellar atrophy on MRI, no. (%)* | 12/20 (60%) | 4/9 (44%) |

No.: number, SD: standard deviation, IQR: Interquartile Range, MRI: magnetic resonance imaging

We studied the meiotic stability of the GAA repeat expansion in a multigenerational French family. The transmission of an expanded (GAA)_467_ allele resulted in expansion in the female germline in two meiotic events (expansion from 467 triplets to 492 and 510 triplets) (Figure 4A: lanes 6,7, and 8). These results are consistent with previous reports showing further expansion of expanded alleles in the female germline^5,6^. In keeping with the transmission of a larger allele in the offspring, clinical anticipation was observed in this family. Both offspring developed episodic ataxia and marked alcohol intolerance more than 10 years earlier than their mother.

## Discussion

The recent discovery of dominantly inherited GAA repeat expansions in *FGF14*^5,6^ as a common cause of LOCA highlights the need for developing an accessible and standardized diagnostic protocol. To address this need, we validated a strategy to genotype the *FGF14* GAA repeat expansion that can easily be implemented in clinical settings. This strategy does not rely on long-read sequencing, a technology that is currently not routinely available in clinical laboratories. We used this approach in a cohort of 53 French patients and confirmed the genetic diagnosis in nine patients and two of their relatives.

This step-wise approach was designed to allow for high-throughput screening while not losing sensitivity to detect expansions. This is a particularly desirable point given the large number of patients with unsolved LOCA who are likely to undergo testing for the *FGF14* GAA repeat expansion. The screening process will end after the second step for the majority of samples, as can been seen in Figure 1. There are, however, a limited number of samples that will need to undergo additional testing, namely gel electrophoresis and Sanger sequencing.

Despite being less automatable and more labor-intensive, these tests are necessary to ensure accurate genotyping given the high degree of length and sequence polymorphism of the *FGF14* repeat locus in the general population^5^. Although local policies can vary with regard to reporting, suggested items to be included in the report are presented in Table S3, depending upon the reason for referral.

We also recommend using bidirectional RP-PCRs targeting both ends of the repeat locus during the second step to interrogate the sequence motif over the entire length of larger expansions, which cannot otherwise be achieved with a single RP-PCR. This further allows for a comprehensive assessment of any potential sequence interruptions and polymorphisms at both ends of the locus. While sequence interruptions are relatively common in Friedreich ataxia, an autosomal recessive disorder caused by a GAA repeat expansion in the *FXN* gene^14^, their frequency and impact on disease expression remain to be established in GAA-*FGF14* ataxia. Current data support a pathogenic threshold for GAA-*FGF14* ataxia of (GAA)_≥250_ uninterrupted repeat units^5,6^. Pending additional data on the effect of interruptions on the pathogenicity of *FGF14* expansions, we suggest that alleles with a minimum repeat tract of 250 uninterrupted triplets be considered pathogenic.

Applying this strategy to a cohort of 53 French patients with unsolved LOCA, we identified nine patients (17%) who carried at least one (GAA)_≥250_ expansion as well as two of their affected relatives. The frequency of the *FGF14* GAA expansion in our cohort was similar to that of other reported cohorts of European descent^5,6^. Patients from the French Canadian and French cohorts had a similar phenotype that included frequent nystagmus and episodic symptoms at onset followed by a slowly evolving pan-cerebellar syndrome at an average age of 59 years in the French Canadian cohort and 66 years in the French cohort. A substantial proportion of patients also reported marked alcohol intolerance that could precede development of ataxia by a number of years in some patients. GAA-*FGF14* ataxia is phenotypically similar to SCA6^15^ and the first late-onset episodic ataxia described thus far. Other genetically defined episodic ataxias, such as episodic ataxia type 1 or 2, often manifest in childhood^16–18^. The observation of a cerebellar syndrome of late onset associated with episodic symptoms, downbeat nystagmus and alcohol intolerance should prompt testing for the *FGF14* GAA expansion. As with other repeat expansion disorders^19^, we found an inverse correlation between the size of the expansion and the age at onset in the French Canadian cohort. However, we did not observe such correlation in the French cohort, likely as a result of the small cohort size.

We found expansion in the female germline across two meiotic events in an affected mother carrying a (GAA)_467_. Clinical anticipation was observed in the offspring who inherited a (GAA)_492_ and (GAA)_510_ allele; both developed episodic ataxia more than 10 years earlier than their mother. These results further highlight the instability of the *FGF14* repeat locus upon meiotic transmission.

In conclusion, the recent description of GAA-*FGF14* ataxia as a common cause of LOCA highlighted the need to develop a reliable diagnostic test for this novel condition. To address this need, we developed and validated an approach to diagnose the *FGF14* GAA repeat expansion that is reproducible, easy to implement in clinical settings and does not rely on long-read sequencing. We implemented this protocol in a clinical laboratory and showed its successful application in a cohort of patients with unsolved LOCA.

## Data Availability

The data analyzed in this study can be accessed upon reasonable request to the corresponding authors.

## Acknowledgments

The authors thank the families for their participation in this project.

This work was supported by the Fondation Groupe Monaco and the Montreal General Hospital Foundation (grant PT79418). The funders had no role in the conduct of this study. D.P. holds a Fellowship award from the Canadian Institutes of Health Research (CIHR).

## Author Contributions

Conceptualization: C.B., D.P., B.B., M.R.; Data curation: C.B., D.P., V.R., G.C., M.W., L.L., S.F., M.D., A.G., I.B., F.W., F.G., C.R., S.C., M.B., C.P., N.D., M.J.D., F.E., M.F.R., A.H., R.L.P., M.S, H.H., M.C.D., S.Z., B.B., M.R.; Formal analysis: C.B., D.P., V.R., G.C., M.W., F.B., F.G., C.R., S.C., M.B., C.P., N.D., M.J.D, F.E., M.F.R., M.C.D, S.Z., B.B., M.R.; Methodology: C.B., D.P., V.R., G.C., M.W., M.J.D., M.S., M.C.D., S.Z., B.B., M.R.; Supervision: C.B., S.Z., B.B., M.R.; Writing-original draft: C.B., D.P., V.R., G.C.; Writing-review & editing: C.B., D.P., V.R., G.C., M.W., L.L., S.F., M.D., A.G., I.B., F.W., F.G., C.R., S.C., M.B., C.P., N.D., M.J.D., F.E., M.F.R., A.H., R.L.P., M.S, H.H., M.C.D., S.Z., B.B., M.R.

## Ethics Declaration

All participants provided written informed consent. This study was conducted in accordance with the Declaration of Helsinki. The institutional review board of the McGill University Health Centre (MPE-CUSM-15-915) and of the Centre Hospitalier Régional Universitaire de Nancy (2020PI220) gave ethical approval for this work.

## Conflict of Interest

Matthis Synofzik has received consultancy honoraria from Janssen, Ionis, Orphazyme, Servier, Reata, GenOrph, and AviadoBio, all unrelated to the present manuscript.

Stephan Zuchner is consultant on drug targets for Aeglea BioTherapeutics and consultant on clinical trial design for Applied Therapeutics, all of them unrelated to the work in the present manuscript.

Other authors have no relevant financial interests to disclose.

## Notes

### Author Declarations

The institutional review board of the McGill University Health Centre (MPE-CUSM-15-915) and of the Centre Hospitalier Regional Universitaire de Nancy (2020PI220) gave ethical approval for this work.

