## Supplementary Appendix for "Optimized testing strategy for the diagnosis of GAA-*FGF14* ataxia"

**Affiliations**

Dr. Céline Bonnet

Laboratoire de génétique médicale, Hôpitaux de Brabois - CHRU de Nancy

Rue du Morvan, 54500 Vandoeuvre-lès-Nancy FRANCE

Dr. Mathilde Renaud

Service de génétique Clinique, Hôpitaux de Brabois - CHRU de Nancy

Rue du Morvan, 54500 Vandoeuvre-lès-Nancy FRANCE

### SUPPLEMENTARY FIGURES

**Figure S1: Testing strategy for the diagnosis of GAA-*FGF14* ataxia**

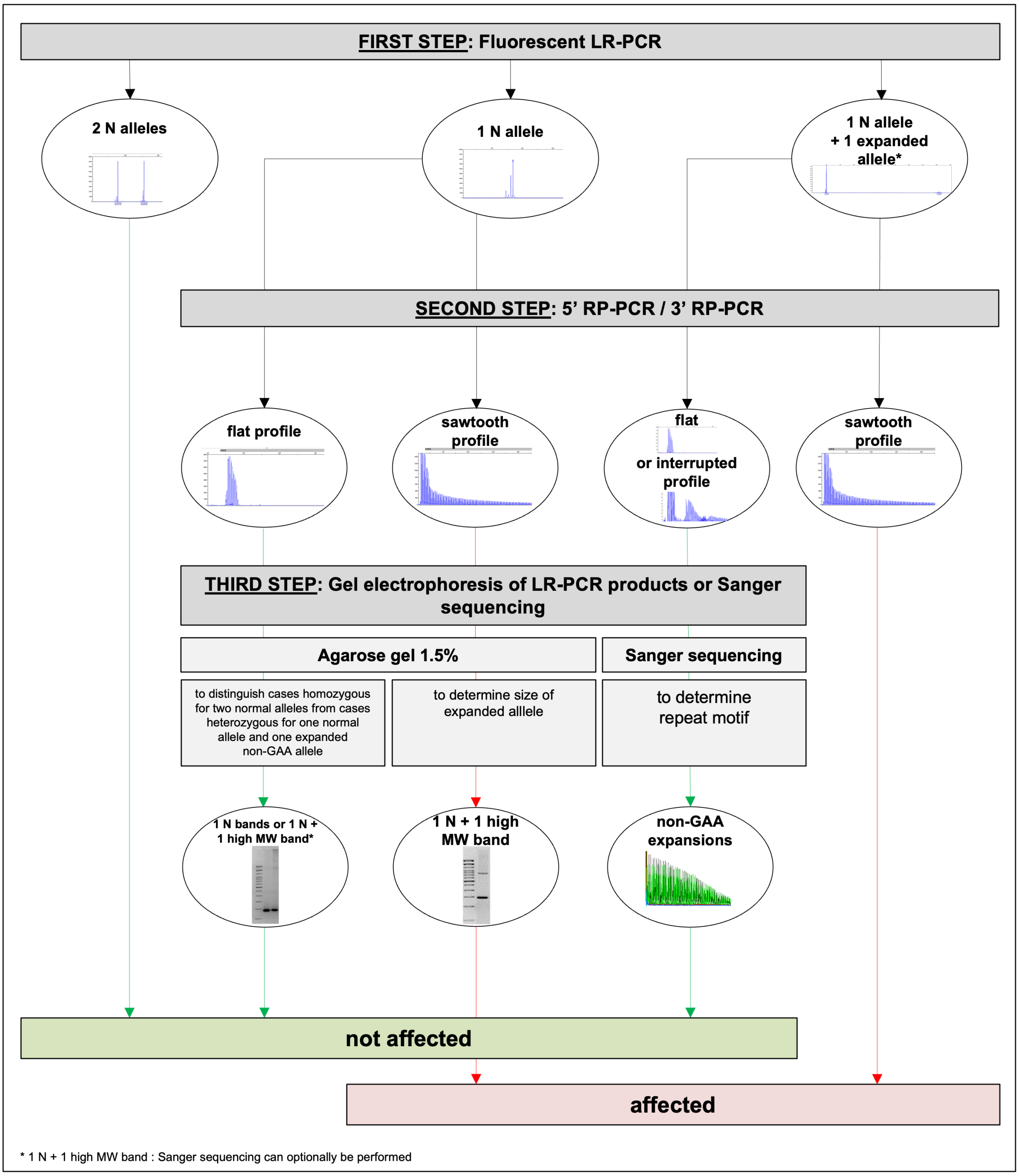


Normal alleles have $(GAA)_{<250}$ repeats and expanded alleles have $(GAA)_{\geq 250}$ repeats.

*Two expanded alleles are possible

Legend: N allele, normal allele <250 repeat units; LR-PCR, long-range polymerase chain reaction; MW, molecular weight.

**Figure S2:** ***FGF14*** **allele size estimates by long-read nanopore sequencing and agarose gel electrophoresis.**

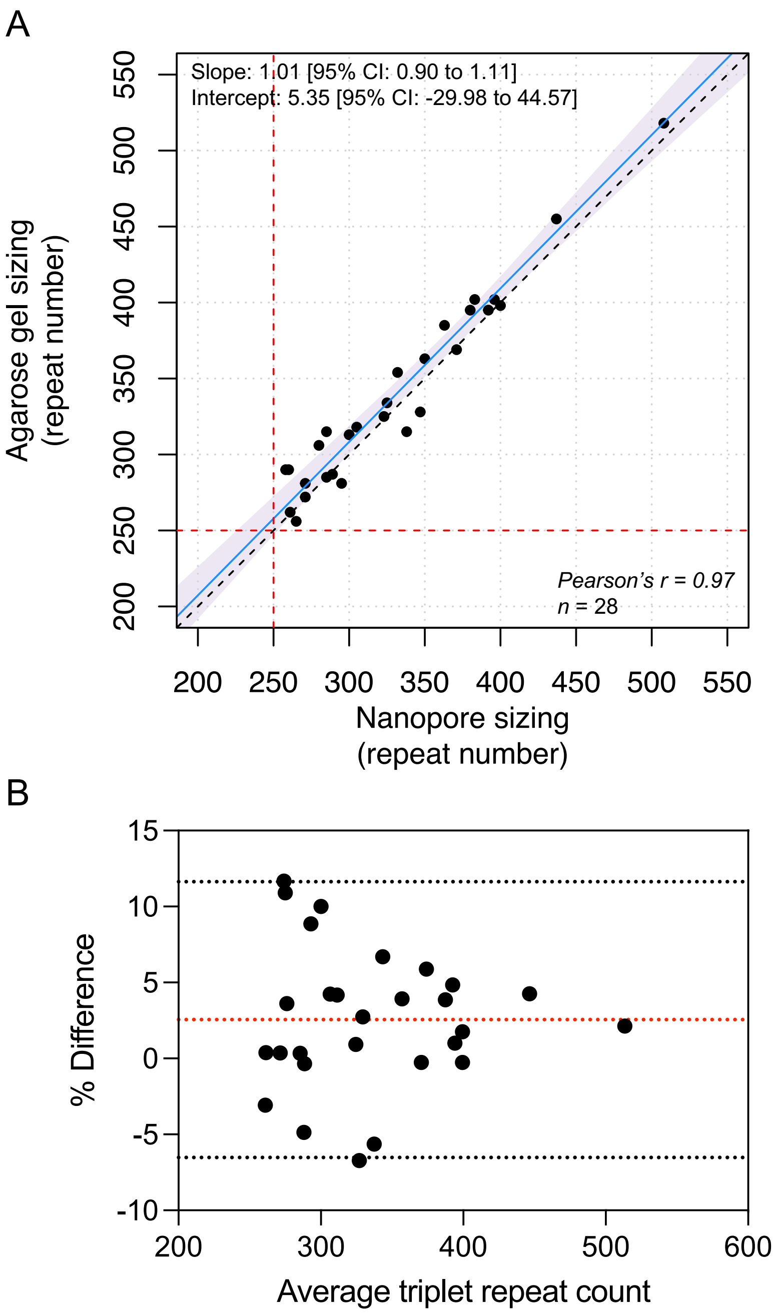


(A) Passing-Bablok regression (blue line) with 95% confidence interval (shaded blue area) for allele size measured by nanopore sequencing and gel electrophoresis. The dashed black line show the identity line and the dashed red lines show the pathogenic threshold of $(GAA)_{\geq 250}$ repeats. (B) Bland-Altman plot shows the percentage difference between size estimates measured by targeted nanopore sequencing and gel electrophoresis as a function of the average of the two measurements for each sample. The dashed red lines show the mean bias between two techniques and the dashed gray lines show the limits of agreement, defined as the mean percentage difference ± 1.96SD.

### SUPPLEMENTARY TABLES

#### Table S1: Experimental conditions used for long-range PCR amplification and RP-PCRs

*Long-range PCR protocols*

| Reagents | Primers | Cycling conditions |
|---|---|---|
| 1. Phusion Flash High-Fidelity PCR Master Mix 2X (Thermo-Fisher)<br>2. Primers 1μM<br>3. gDNA 40ng | **LR-PCR**<br>F: TGCAAATGAAGGAAAACTCTT<br>R: CAATGATGAATTAAGCAGTTCC<br><br>**fLR-PCR**<br>F: 6-FAM-TGCAAATGAAGGAAAACTCTT<br>R: CAATGATGAATTAAGCAGTTCC | 98°C x 3 min<br>[98°C x 10 sec<br>65°C x 15 sec – Each 2 cycles decreasing by 1°C<br>72°C x 3 min] x 12 cycles<br><br>[98°C x 10 sec<br>59°C x 15 sec<br>72°C x 3 min] x 20 cycles<br><br>72°C x 5 min |

*RP-PCR protocols*

| Reagents | Primers | Cycling conditions |
|---|---|---|
| 1. Phusion Flash High-Fidelity PCR Master Mix 2X (Thermo-Fisher)<br>2. F primer 1μM<br>3. R primer 0.1μM<br>4. M13-FAM 1μM<br>5. gDNA 80ng | **3'RP-PCR**<br>F: TGCCCACATAGAGCTTAGTCT<br>R: CACGACGTTGTAAAACGAC-GAAGAAGAAGAAGAAGAAGAA<br>M13-FAM: 6-FAM-CACGACGTTGTAAAACGAC<br><br>**5'RP-PCR**<br>F: TGCAAATGAAGGAAAACTCTT<br>R: CACGACGTTGTAAAACGAC-TTCTTCTTCTTCTTCTTCTTC<br>M13-FAM: 6-FAM-CACGACGTTGTAAAACGAC | 98°C x 3 min<br><br>[98°C x 10 sec<br>65°C x 15 sec<br>72°C x 1 min] x 35 cycles<br><br>72°C x 5 min |

F, forward; gDNA, genomic DNA; PCR, polymerase chain reaction; R, reverse

**Table S2:** Comparison of sizing estimates by fluorescent LR-PCR (with and without correction) and targeted nanopore sequencing.

| **Nanopore sizing** | **fluorescent LR-PCR sizing** | | | | |
|---|---|---|---|---|---|
| **repeat number [size bp]** | **[size bp]** | **repeat number calculated** | | **corrected** | |
| 258 | 876.90 | 242.30 | -6% | 266.02 | 3% |
| 260 | 873.71 | 241.24 | -7% | 264.83 | 2% |
| 261 | 879.92 | 243.31 | -7% | 267.14 | 2% |
| 265 | 874.36 | 241.45 | -9% | 265.07 | 0% |
| 271 | 900.00 | 250.00 | -8% | 274.63 | 1% |
| 271 | 905.57 | 251.86 | -7% | 276.70 | 2% |
| 280 | 921.94 | 257.31 | -8% | 282.81 | 1% |
| 285 | 955.35 | 268.45 | -6% | 295.26 | 4% |
| 285 | 929.72 | 259.91 | -9% | 285.71 | 0% |
| 289 | 952.90 | 267.63 | -7% | 294.35 | 2% |
| 295 | 936.54 | 262.18 | -11% | 288.25 | -2% |
| 300 | 1035.55 | 295.18 | -2% | 325.15 | 8% |
| 305 | 1003.79 | 284.60 | -7% | 313.31 | 3% |
| 323 | 1039.49 | 296.50 | -8% | 326.62 | 1% |
| 325 | 1057.28 | 302.43 | -7% | 333.25 | 3% |
| 332 | 1060.24 | 303.41 | -9% | 334.36 | 1% |
| 338 | 1041.80 | 297.27 | -12% | 327.48 | -3% |
| 347 | 1062.12 | 304.04 | -12% | 335.06 | -3% |
| 350 | 1121.00 | 323.67 | -8% | 357.01 | 2% |
| 363 | 1173.86 | 341.29 | -6% | 376.71 | 4% |
| 371 | 1153.54 | 334.51 | -10% | 369.13 | -1% |
| 380 | 1215.20 | 355.07 | -7% | 392.12 | 3% |
| 383 | 1208.65 | 352.88 | -8% | 389.68 | 2% |
| 392 | 1228.55 | 359.52 | -8% | 397.09 | 1% |
| 396 | 1220.51 | 356.84 | -10% | 394.10 | 0% |
| 400 | 1214.87 | 354.96 | -11% | 392.00 | -2% |

Slope: 2.6827; y-intercept: 163.2416

Internal control: $(GAA)_{10}$ : 180,32 bp; $(GAA)_{258}$ :876.90 bp; $(GAA)_{323}$ :1039.69 bp; $(GAA)_{400}$ :1214.87 bp.

**Table S3:** Suggested items to be included in the report

| Range: number of repeats | Reason for referral | |
|---|---|---|
| | **Diagnostic testing** | **Carrier testing** |
| **Normal**<br>-Allele 1 <250 repeat units<br>-Allele 2 <250 repeat units | The diagnosis of GAA-*FGF14* (SCA27B, late-onset) ataxia is excluded | Patient is not a carrier of a *FGF14* GAA repeat expansion |
| **Non-GAA expansion: ≥250 repeat units**<br>-Allele 1 <250 repeat units<br>-Allele 2 ≥250 non-GAA repeat units | The diagnosis of GAA-*FGF14* ataxia (SCA27B, late-onset) is excluded | - |
| **GAA expansion: ≥250 repeat units**<br>-Allele 1 <250 repeat units<br>-Allele 2 ≥250 GAA repeat units<br>or<br>-Allele 1 ≥250 GAA repeat units<br>-Allele 2 ≥250 GAA repeat units | The diagnosis of GAA-*FGF14* ataxia (SCA27B, late-onset) is confirmed.<br>The report should mention the indication of genetic counselling for the family of the patient (offering confirmatory carrier testing) | The patient is a carrier of a *FGF14* GAA repeat expansion |
